# SUBSTANCE USE AS A PREDICTOR OF RISKY SEXUAL BEHAVIOR AND STI RISK AMONG URBAN YOUTH IN GHANA

**DOI:** 10.64898/2026.06.18.26355934

**Authors:** Osei-Mensah Rockson, Faustina Hayford Blankson

**Author notes:** Corresponding author (FHB).

## Abstract

**Background:** Substance use and risky sexual behavior (RSB) are intertwined public health threats that drive sexually transmitted infection (STI) transmission among young people. Despite escalating drug misuse in urban Ghana, integrated evidence on the drug-RSB-STI triad remains scarce, particularly for opioid substances such as tramadol and for the full spectrum of RSB practices.

**Objective:** This study examined the prevalence of substance use and RSBs, quantified associations between substance use and RSB, and documented the RSB-STI relationship among youth aged 18 to 35 years in Ablekuma North Municipality, Greater Accra, Ghana.

**Methods:** A cross-sectional quantitative design was employed with 383 participants recruited via stratified random sampling across 14 electoral areas. Validated instruments assessed substance use, RSB, and self-reported STI symptoms. Pearson correlation, hierarchical multiple regression, and chi-square tests of independence with odds ratios constituted the primary analytic approaches.

**Results:** Alcohol was the most prevalent substance (51.7%), followed by aphrodisiacs (26.4%), shisha (19.6%), cigarettes (15.9%), and tramadol (14.6%). RSBs were widespread: 50.9% reported condomless sex, 47.5% unplanned sex, 45.2% sex without pregnancy protection, and 19.6% transactional sex. Gonorrhea (18.8%) and chlamydia (14.4%) were the most prevalent STI symptoms; HIV positivity stood at 15.6% among those tested, with 59.4% unaware of their status. Drug abuse strongly predicted RSB (β = .727, p < .001; ΔR² = .409). Tramadol users were up to 49.6 times more likely to have sex with casual partners and 31.4 times more likely to engage in transactional sex. RSBs were consistently and significantly associated with HIV, gonorrhea, syphilis, chlamydia, and herpes (all p < .001).

**Conclusion:** Tramadol is an underrecognized and high-magnitude driver of sexual risk in urban Ghana. The high prevalence of RSBs alongside low STI testing and treatment uptake constitutes a compounding public health crisis. Integrated substance-use and sexual health interventions are urgently required.

## INTRODUCTION

Globally, an estimated 296 million people used illicit substances in 2021, representing a 23% increase from 240 million in 2011, with sub-Saharan Africa projected to sustain a 40% increase in drug consumption by 2030.^1,2^ Among young people, substance use is not merely a health burden in its own right; it operates as a proximal, pharmacological driver of RSB and, through RSB, of STI transmission. These three phenomena constitute an epidemiological triad that disproportionately concentrates on low-and middle-income urban settings where poverty, unemployment, peer influence, and drug accessibility converge.

In Ghana, approximately 70% of drug users are under 35 years of age.^3^ The substance landscape has shifted considerably in recent years, with tramadol (an opioid analgesic subject to widespread non-medical use) emerging as a drug of major concern alongside alcohol and cannabis.^4,5^ Concurrently, Ghana’s HIV prevalence among youth aged 15 to 24 stands at 1.2%, while bacterial STIs such as gonorrhea and syphilis remain substantially underreported due to limited testing infrastructure and low health-seeking behaviour.^6,7^ Studies in Greater Accra document high rates of condomless sex, multiple sexual partnerships, and transactional sex among urban youth,^8,9^ each of which intersects with both substance use and elevated STI risk. Yet these three phenomena have rarely been examined together in a single community-based study with disaggregated, substance-specific analyses.

The pharmacological mechanism linking substance use to RSB is well established. Psychoactive substances including alcohol, tramadol, cannabis, and shisha impair prefrontal cortical function, the primary neurological substrate of impulse control, risk evaluation, and inhibitory regulation.^10^ This disinhibition reduces the motivational salience of negative health consequences and increases impulsive engagement in sexual risk-taking.^11,12^ Critically, tramadol’s dopaminergic and serotonergic pharmacodynamic profile makes it particularly potent in altering reward-driven decision-making and lowering social inhibitions around sexual behaviour.^4,5^ Despite this, tramadol’s downstream sexual health consequences remain poorly characterized in the empirical literature relative to alcohol.

A further dimension of this epidemic is the prevalence and patterning of RSBs themselves. Prior Ghanaian studies have documented individual risk behaviours,^8,13^ but comprehensive prevalence estimates covering the full RSB spectrum (unplanned sex, regretted sex, multiple partnerships, condomless sex, sex without pregnancy protection, casual sex, substance use during sex, and transactional sex) within a single community-based sample are lacking. This gap is consequential: the magnitude and distribution of RSBs in a population determine the network-level risk of STI transmission, and interventions must be calibrated to the specific behavioral profile of the target population rather than to a generalized risk profile.^14^

This study addresses four objectives: (1) to determine the prevalence of substance use, RSBs, and STI symptoms among youth aged 18 to 35 in Ablekuma North Municipality, Greater Accra; (2) to examine the association between substance use and RSB, disaggregated by substance type; (3) to quantify the associations between specific RSBs and STI outcomes; and (4) to contextualize these findings within the broader sub-Saharan African literature on the drug-RSB-STI pathway. The findings contribute novel, disaggregated evidence that positions tramadol as a priority substance for integrated prevention programing in urban Ghana.

### Theoretical Framework

This study draws on the Health Belief Model (HBM) and the Theory of Planned Behavior (TPB) as complementary frameworks for understanding substance use, RSB, and STI vulnerability. The HBM posits that health behavior is shaped by perceived susceptibility to a health threat, perceived severity of its consequences, perceived benefits of protective action, and barriers to taking such action.^15^ In this context, the HBM helps explain why substance-using youth may persist in RSB even when they are cognitively aware of STI risks: low perceived susceptibility, or fatalistic resignation rooted in high local STI prevalence, can override knowledge-based protective intent.^16^ High unemployment and economic precarity (documented in this population) further erode perceived agency, weakening the cues to action that the HBM identifies as necessary for behavior change.

The TPB holds that behavior is determined by intentions, which are shaped by three constructs: attitudes toward the behavior, subjective norms reflecting perceived social pressure, and perceived behavioral control.^15^ Peer environments that normalize drug use in Ablekuma North simultaneously normalize risky sexual practices,^17,18^ producing a convergent normative pressure that weakens individual protective intentions. Both frameworks, however, have a critical limitation in the context of substance use: neither accounts adequately for the acute pharmacological disinhibition that psychoactive substances produce, a mechanism capable of overriding cognitive risk appraisal processes entirely.^10,11^ The study’s conceptual framework therefore positions substance abuse as the primary independent variable directly driving RSB, which in turn drives STI transmission, with HBM and TPB constructs framing the psychosocial context within which this pharmacological pathway operates. Demographic variables (age, sex, education, employment) are incorporated as confounders.

## METHODS

### Study Design, Setting, and Participants

A cross-sectional quantitative design was employed, appropriate for estimating the prevalence of health behaviors and quantifying associations between exposures and outcomes at a defined point in time.^19^ The study was conducted in Ablekuma North Municipality, Greater Accra, which had a 2021 census population of 159,208.^20^ The municipality is characterized by rapid urbanization, high youth density, and increasing accessibility to psychoactive substances, making it an epidemiologically relevant setting.^21^

Sample size was calculated using the Yamane (1967) formula (n = N / 1 + N[e]²), yielding a minimum sample of 400 (N = 159,208; e = 0.05), inflated to 440 to account for non-response. A stratified random sampling technique divided the municipality into 14 strata corresponding to its electoral areas, from which participants were recruited proportionally, ensuring equitable geographic and demographic representation.^22^ Eligible participants were youth aged 18 to 35 years, residing in the municipality at the time of data collection. A total of 383 valid responses were obtained (response rate: 87.0%). Participants were recruited from community centers, religious venues, markets, and social spaces by trained research assistants.

### Measures

#### Substance use

Assessed over 6 months using a researcher-developed six-item scale covering alcohol, cigarettes, marijuana, tramadol, shisha, and aphrodisiacs (Cronbach’s α = .927). Each item was scored on a five-point Likert scale (0 = never to 4 = very often), generating a composite drug abuse score (range: 0 to 30). Higher scores indicated greater substance use frequency.

#### Risky sexual behavior

Measured using eight items adapted from the validated Sexual Risk Survey (SRS; Turchik and Garske, 2009; α = .929), capturing frequency of eight RSBs in the preceding six months: unplanned sex, regretted sex, multiple sexual partners, condomless sex, sex without pregnancy protection, sex with casual partners, substance use before or during sex, and transactional sex. Items were scored ordinally (0 to 4), generating a total RSB score (range: 0 to 32).

#### STI outcomes

Assessed through self-reported symptoms of HIV, gonorrhea, syphilis, chlamydia, and herpes over the preceding six months, alongside HIV testing history, treatment-seeking behavior, and partner disclosure. HIV status was additionally captured from those who had been tested.

### Statistical Analysis

Descriptive statistics (frequencies, percentages) summarized participant characteristics, substance use, RSB prevalence, and STI outcomes. For the association between drug abuse and RSB, Pearson’s r quantified the bivariate relationship. Hierarchical multiple regression assessed the independent predictive contribution of drug abuse to RSB: Model 1 entered demographic controls (sex, age group, education level, employment status, religious affiliation); Model 2 added the composite drug abuse score. For chi-square analyses, substance use items were dichotomized (0 = never; 1 = any use), and RSB items were similarly dichotomized (0 = no engagement; 1 = any engagement), except multiple sexual partners, where two or more partners constituted engagement. Odds ratios (OR) with 95% confidence intervals (CI) quantified association magnitudes. All analyses used SPSS version 26; significance was set at p < .05.

### Ethical Considerations

Ethical approval was granted by the Noguchi Memorial Institute for Medical Research Institutional Review Board (NMIMR-IRB; approval number: NMIMR IRB CPN: 067/24-25). All participants provided written informed consent. Participation was voluntary; participants were informed of their right to withdraw without consequence. Data were anonymized, password-protected, and stored securely. Sensitive subject matter was addressed using culturally appropriate, neutral language.

## RESULTS

### Sample Characteristics, Prevalence of Substance Use, Risky Sexual Behavior, and STI Outcomes

Of 383 participants, 50.1% were male, 48.3% female, and 1.6% identified as another gender. Most were aged 18 to 24 years (53.8%), had attained tertiary education (61.6%), and were unemployed (42.0%). Christianity was the dominant religious affiliation (82.0%). The striking contrast between high educational attainment and high unemployment reflects a structural vulnerability consistent with increased risk of both substance use and transactional sex in this age group.^23^

Table 1 presents substance use prevalence, RSB prevalence, and STI outcomes as an integrated descriptive profile of the sample. Alcohol was the most used substance (51.7%), followed by aphrodisiacs (26.4%), shisha (19.6%), cigarettes (15.9%), and tramadol (14.6%). The motivational landscape is notable: sexual performance enhancement was cited as a reason for tramadol use by 3.9% of the sample, and for alcohol use by 14.4%, indicating that for a meaningful subset of participants, the drug-RSB relationship is not incidental but partly intentional.^4,5^

**Table 1.** Integrated prevalence profile: substance use, risky sexual behaviors, and STI outcomes among youth in Ablekuma North, Ghana (N = 383).

| Variable | n | % | Contextual note |
| --- | --- | --- | --- |
| **SUBSTANCE USE (past 6 months)** |  |  |  |
| Alcohol | 198 | 51.7 | Fun 20.6%; sexual performance 14.4%; stress 3.1% |
| Aphrodisiacs | 101 | 26.4 | Cream/spray 11.2%; herbal-alcohol base 7.0% |
| Shisha | 75 | 19.6 | Fun 11.5%; sexual performance 3.4%; peer influence 2.6% |
| Cigarettes | 61 | 15.9 | Sexual performance 6.8%; stress 4.2%; peer influence 2.9% |
| Marijuana | 56 | 14.6 | Stress 7.3%; sexual performance 4.4%; fun 1.3% |
| Tramadol (non-prescribed) | 56 | 14.6 | Stress 5.7%; sexual performance 3.9%; peer influence 2.6% |
| **RISKY SEXUAL BEHAVIOUR (past 6 months; any engagement)** |  |  |  |
| Condomless sex | 195 | 50.9 | Highest RSB prevalence in sample |
| Unplanned sex | 182 | 47.5 |  |
| Sex without pregnancy protection | 173 | 45.2 |  |
| Regretted sex | 164 | 42.8 | Suggests impulsive or substance-influenced decision-making |
| Sex with casual partners | 135 | 35.2 | 'Friends with benefits' or non-romantic partners |
| Substance use before or during sex | 98 | 25.6 | Deliberate use to facilitate sexual activity |
| Multiple sexual partners (2 or more) | 113 | 29.5 |  |
| Transactional sex | 75 | 19.6 | Exchange for money, academic grades, or favors |
| **STI OUTCOMES (past 6 months)** |  |  |  |
| HIV tested | 98 | 25.6 | 74.4% had never been tested |
| HIV positive (of those tested) | 59 | 15.6 | 59.4% unaware of their status |
| Gonorrhea symptoms | 72 | 18.8 | Most prevalent STI symptom |
| Chlamydia symptoms | 55 | 14.4 |  |
| Syphilis symptoms | 41 | 10.7 |  |
| Herpes symptoms | 38 | 9.9 |  |
| Sought treatment for STI symptoms | 83 | 21.7 | 78.3% did not seek treatment despite symptoms |
| Disclosed symptoms to sexual partner | 50 | 13.1 | 86.9% did not disclose; risk of onward transmission |

*Note.* RSB = risky sexual behavior. STI = sexually transmitted infection. Substance use motivations reported as percentage of total sample. RSB prevalence reflects participants reporting at least one episode in the preceding six months. Multiple sexual partners are defined as two or more partners.

RSBs were highly prevalent across the sample. Condomless sex was reported by 50.9% of participants, unplanned sex by 47.5%, sex without pregnancy protection by 45.2%, regretted sex by 42.8%, sex with casual partners by 35.2%, substance use during sex by 25.6%, and transactional sex by 19.6%. These rates are comparable to those documented among high-risk groups including female sex workers and men who have sex with men in Ghanaian urban settings,^24,25^ and markedly exceed population-level estimates from nationally representative surveys,^26^ underscoring the elevated risk profile of this sample.

STI burden was substantial. Gonorrhea symptoms were the most prevalent (18.8%), followed by chlamydia (14.4%), syphilis (10.7%), and herpes (9.9%). These rates substantially exceed the global average below 5% in low-risk groups and are consistent with data from high-risk sub-Saharan African populations.^27^ HIV positivity stood at 15.6% among those tested, though only 25.6% of participants had ever been tested for HIV, and 59.4% were unaware of their status. Healthcare-seeking behavior was critically low: only 21.7% sought treatment for STI symptoms, and only 13.1% disclosed symptoms to sexual partners. Together, these figures describe a population in which infection is widespread, diagnosis is rare, treatment is uncommon, and partner notification is the exception rather than the rule.

### Association Between Substance Use and Risky Sexual Behavior

Pearson correlation revealed a strong, positive, and statistically significant bivariate association between drug abuse and RSB (r = .743, p < .001). Table 2 presents the hierarchical regression results. Model 1 (demographics only) was statistically significant (F [5, 377] = 14.88, p < .001; R²= .165), with age group (β = .354, p < .001), education level (β = −.222, p < .001), and employment status (β = .209, p < .001) as the significant predictors. Religious affiliation and sex were non-significant. The introduction of drug abuse in Model 2 yielded a significant improvement in model fit (F [6, 376] = 84.49, p < .001; R² = .574; ΔR² = .409), with drug abuse emerging as the dominant predictor (β = .727, p < .001; B = 1.311, 95% CI [1.18, 1.45]). Demographic predictors that were significant in Model 1 (including education level and employment status) became non-significant once drug abuse entered the model, indicating that a substantial portion of their apparent influence on RSB may be mediated through or confounded by substance use. All regression assumptions were met: tolerance = 0.775; VIF = 1.290; Durbin-Watson = 1.92.

**Table 2.**
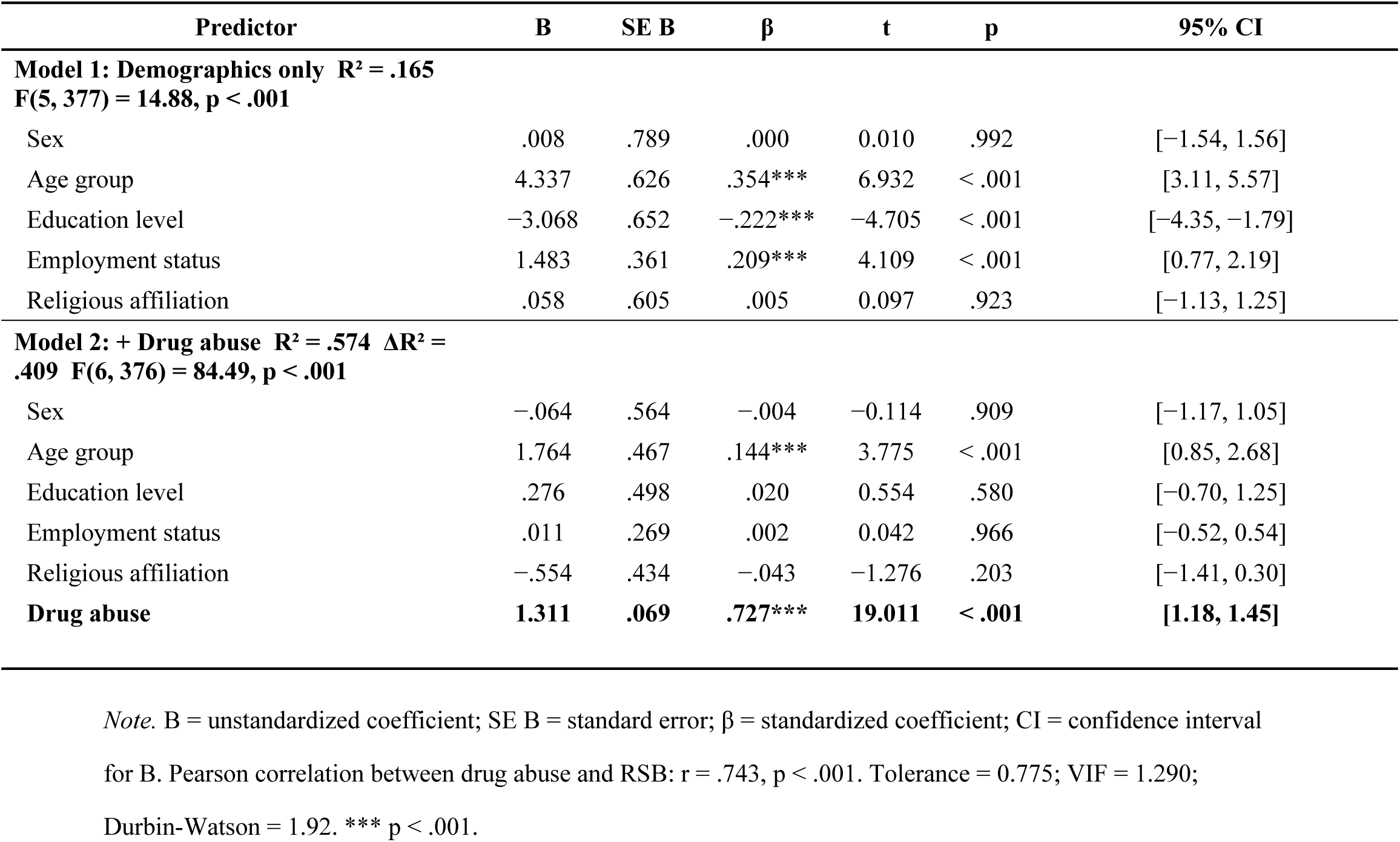
Hierarchical multiple regression predicting risky sexual behavior: demographics-only model (Model 1) and demographics plus drug abuse model (Model 2) (N = 383).

| Predictor | B | SE B | $\beta$ | t | p | 95% CI |
| --- | --- | --- | --- | --- | --- | --- |
| **Model 1: Demographics only $R^2 = .165$**<br>**$F(5, 377) = 14.88, p < .001$** |  |  |  |  |  |  |
| Sex | .008 | .789 | .000 | 0.010 | .992 | [-1.54, 1.56] |
| Age group | 4.337 | .626 | .354*** | 6.932 | < .001 | [3.11, 5.57] |
| Education level | -3.068 | .652 | -.222*** | -4.705 | < .001 | [-4.35, -1.79] |
| Employment status | 1.483 | .361 | .209*** | 4.109 | < .001 | [0.77, 2.19] |
| Religious affiliation | .058 | .605 | .005 | 0.097 | .923 | [-1.13, 1.25] |
| **Model 2: + Drug abuse $R^2 = .574$ $\Delta R^2 = .409$ $F(6, 376) = 84.49, p < .001$** |  |  |  |  |  |  |
| Sex | -.064 | .564 | -.004 | -0.114 | .909 | [-1.17, 1.05] |
| Age group | 1.764 | .467 | .144*** | 3.775 | < .001 | [0.85, 2.68] |
| Education level | .276 | .498 | .020 | 0.554 | .580 | [-0.70, 1.25] |
| Employment status | .011 | .269 | .002 | 0.042 | .966 | [-0.52, 0.54] |
| Religious affiliation | -.554 | .434 | -.043 | -1.276 | .203 | [-1.41, 0.30] |
| **Drug abuse** | **1.311** | **.069** | **.727\*\*\*** | **19.011** | **< .001** | **[1.18, 1.45]** |

Note. B = unstandardized coefficient; SE B = standard error; $\beta$ = standardized coefficient; CI = confidence interval for B. Pearson correlation between drug abuse and RSB: $r = .743, p < .001$ . Tolerance = 0.775; VIF = 1.290; Durbin-Watson = 1.92. \*\*\* $p < .001$ .

Table 3 presents chi-square associations between specific substances and specific RSBs. All associations were significant at p < .001. Alcohol users were 12.21 times more likely to have multiple sexual partners (50.0% vs. 7.6%) and 13.30 times more likely to engage in transactional sex (34.3% vs. 3.8%). Cigarette users exhibited exceptionally high odds for condomless sex (OR = 81.13; 98.3% vs. 42.1%), casual sex (OR = 59.68), and transactional sex (OR = 84.78). Tramadol users demonstrated similarly extreme risk profiles: 88.9% reported condomless sex (OR = 9.91), 85.2% multiple partnerships (OR = 22.49), 94.4% casual sex (OR = 49.58), and 77.8% transactional sex (OR = 31.39). These odds ratios indicate that disinhibition operates selectively and with greater potency for tramadol and cigarettes than for alcohol in the domain of the most structurally embedded risk behaviors.

**Table 3.**
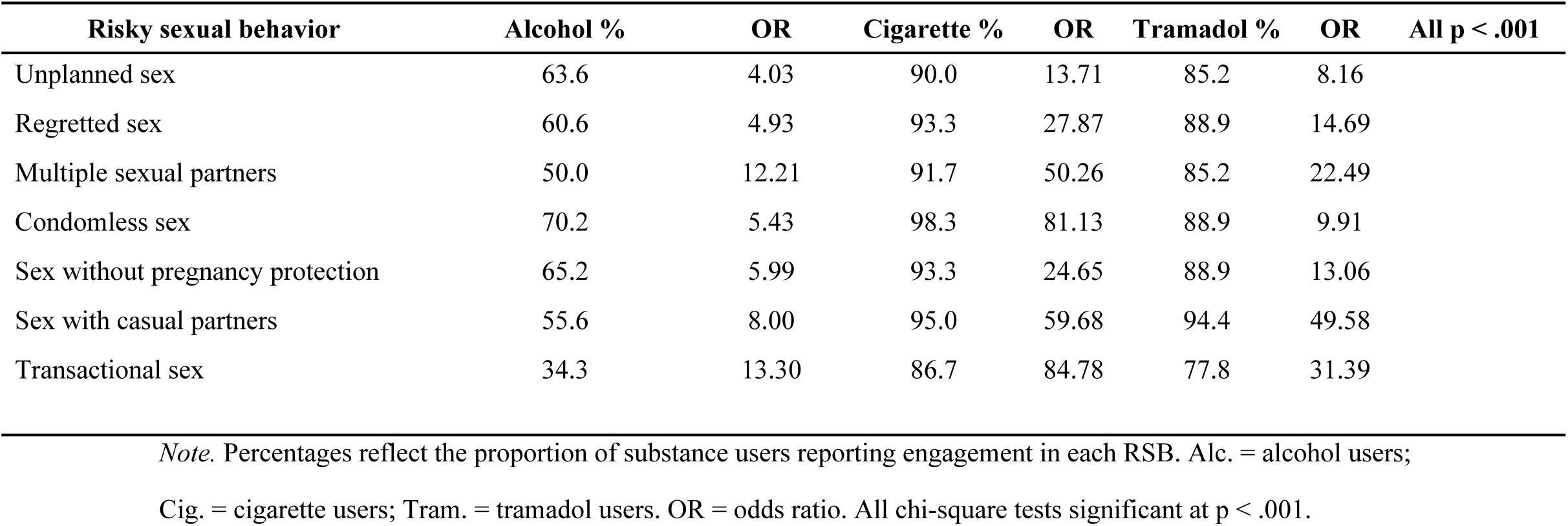
Associations between specific substance use and risky sexual behaviors: chi-square tests of independence (N = 383).

| Risky sexual behavior | Alcohol % | OR | Cigarette % | OR | Tramadol % | OR | All p < .001 |
| --- | --- | --- | --- | --- | --- | --- | --- |
| Unplanned sex | 63.6 | 4.03 | 90.0 | 13.71 | 85.2 | 8.16 |  |
| Regretted sex | 60.6 | 4.93 | 93.3 | 27.87 | 88.9 | 14.69 |  |
| Multiple sexual partners | 50.0 | 12.21 | 91.7 | 50.26 | 85.2 | 22.49 |  |
| Condomless sex | 70.2 | 5.43 | 98.3 | 81.13 | 88.9 | 9.91 |  |
| Sex without pregnancy protection | 65.2 | 5.99 | 93.3 | 24.65 | 88.9 | 13.06 |  |
| Sex with casual partners | 55.6 | 8.00 | 95.0 | 59.68 | 94.4 | 49.58 |  |
| Transactional sex | 34.3 | 13.30 | 86.7 | 84.78 | 77.8 | 31.39 |  |

*Note.* Percentages reflect the proportion of substance users reporting engagement in each RSB. Alc. = alcohol users; Cig. = cigarette users; Tram. = tramadol users. OR = odds ratio. All chi-square tests significant at $p < .001$ .

### Association Between Risky Sexual Behavior and STI Outcomes

Table 4 presents RSB-STI associations. All were statistically significant (p < .001). Multiple sexual partners conferred the highest risk for gonorrhea symptoms (OR = 36.76; 55.8% of those with multiple partners vs. 3.3% without) and elevated odds for all other STIs. Condomless sex was associated with the highest syphilis (OR = 47.62) and herpes (OR = 43.48) odds ratios in the dataset. Transactional sex demonstrated the strongest HIV association (OR = 9.41; 44.6% HIV-positive among those engaging in transactional sex vs. 8.5% among non-engagers) and consistently high odds across all STI outcomes. Alcohol use during sex produced the broadest elevation: 40.2% of those using alcohol during sex were HIV-positive, with syphilis OR reaching 47.62.

**Table 4.** Associations between risky sexual behaviors and STI outcomes: chi-square tests of independence with odds ratios (N = 383).

| Risky sexual behavior | HIV+ % | OR | Gonorrhea % | OR | Syphilis % | OR | Chlamydia % | OR |
| --- | --- | --- | --- | --- | --- | --- | --- | --- |
| Multiple sexual partners | 35.7 | 5.63 | 55.8 | 36.76 | 34.5 | 28.57 | 38.9 | 14.93 |
| Condomless sex | 22.7 | 3.95 | 34.9 | 24.39 | 20.5 | 47.62 | 26.2 | 16.39 |
| Alcohol use during sex | 40.2 | - | 63.3 | 24.39 | 38.8 | 47.62 | 43.9 | 16.39 |
| Transactional sex | 44.6 | 9.41 | 68.0 | 29.41 | 45.3 | 35.71 | 50.7 | 19.30 |

*Note.* Percentages reflect STI outcome prevalence among those engaging in the specified RSB. OR = odds ratio. All chi-square tests significant at $p < .001$ . Gonorrhea. = gonorrhea; Chlamydia = chlamydia. OR for HIV not calculable due to polytomous HIV status variable (positive/negative/unknown); chi-square significant at $p < .001$ .

Mapping these findings onto the RSB prevalence data in Table 1 clarifies the public health arithmetic: with 50.9% of the sample reporting condomless sex and associated syphilis odds of 47.62, and 19.6% reporting transactional sex with HIV odds of 9.41, the proportion of infections attributable to these two behaviors alone is substantial. The low treatment-seeking rate (21.7%) and near-absent partner disclosure (13.1%) mean that most infections are neither treated nor prevented from onward transmission, compounding the epidemic at the network level.

## DISCUSSION

This study provides integrated, disaggregated evidence on the drug-RSB-STI triad in an urban Ghanaian community, with four analytically distinct contributions: the documentation of a high-prevalence RSB landscape that extends well beyond prior single-behavior estimates; the quantification of tramadol as a disproportionately high-risk substance for the most structurally embedded RSBs; the confirmation that substance use explains a uniquely large proportion of RSB variance independent of demographics; and the mapping of a structural RSB-STI risk gradient anchored at transactional sex and alcohol use during intercourse.

### The RSB Landscape: Prevalence and Public Health Significance

The RSB prevalence rates documented in this study (50.9% for condomless sex, 47.5% for unplanned sex, and 19.6% for transactional sex) substantially exceed estimates from school-based and nationally representative surveys in Ghana.^26,28^ They are, however, broadly consistent with rates reported among high-risk groups in urban Ghanaian settings. Dery et al.^24^ documented 45 to 50% condomless sex among female sex workers in Accra, and Malefo et al.^25^ reported 40% condomless sex among men who have sex with men in South Africa. That a community-based sample of youth in Ablekuma North approaches these rates suggests a population whose risk profile overlaps considerably with groups traditionally considered high-risk, rather than a general population sample with discrete high-risk subgroups.

The prevalence of transactional sex (19.6%) is particularly noteworthy. Transactional sex in this context is not a marginal behavior concentrated in a small high-risk subset; it is reported by nearly one in five participants and is embedded in a context of high youth unemployment (42%) and high educational attainment (61.6% tertiary). This paradox (educated but economically excluded) creates a structural demand for transactional sex as an economic coping strategy,^23,29^ and it is precisely this structural driver that makes tramadol and alcohol facilitation of transactional sex so consequential: the substance use lowers the inhibitory threshold for a behavior already under economic pressure. Ranganathan et al.^29^ documented similar dynamics among young women in rural South Africa, where transactional sex was embedded in economic need rather than sex work identity, underscoring the generalization of this structural pathway across sub-Saharan African settings.

The motivational profile of substance use reinforces this interpretation. That, 14.4% of the sample used alcohol specifically to enhance sexual performance, and 3.9% used tramadol for the same purpose, confirms that for a meaningful subset of participants, the drug-RSB relationship is partially intentional rather than purely incidental. This deliberate disinhibition represents a behavioral pathway that awareness-based interventions are unlikely to disrupt, since participants are not unaware of what they are doing, they are making a motivated decision under pharmacological influence.

### Substance Use as the Dominant Predictor of RSB

The finding that drug abuse explained an additional 40.9% of RSB variance beyond all demographic controls (ΔR² = .409; β = .727, p < .001) positions substance use as the primary modifiable determinant of sexual risk-taking in this population. This effect size substantially exceeds those reported in comparable regression analyses in Ghana and sub-Saharan Africa. Doku^14^ reported significant but considerably more modest alcohol-RSB associations among Ghanaian youth, while a meta-analysis by Cho and Yang^30^ found pooled ORs of 1.96 for early sexual initiation and 1.72 for multiple partners attributable to alcohol across African adolescent samples. The considerably stronger effects observed here likely reflect the more elevated risk profile of this community-based urban sample relative to the school-going populations that dominate the prior literature.

The attenuation of demographic predictors (particularly education level and employment status) upon entry of drug abuse into the regression model is analytically significant. It suggests that the apparent protective effect of education on RSB, and the apparent risk-increasing effect of unemployment, are substantially mediated through or confounded by substance use patterns. In other words, educated but unemployed youth may be at risk primarily because unemployment predisposes them to substance use, which in turn drives RSB, rather than because unemployment directly increases sexual risk-taking through some independent pathway. This has a direct implication for intervention: addressing unemployment without addressing substance use may be insufficient to reduce RSB in this population.

The role of polysubstance co-use further amplifies this picture. The clustering of substance use behaviors (95% cigarette-shisha co-use, 86.7% cigarette-marijuana co-use, 83.3% tramadol-shisha co-use) indicates that substance use in this population operates as a behavioral ecology rather than a set of independent exposures.^17^ Single-substance analyses, including the chi-square results presented in Table 3, therefore underestimate the cumulative substance-attributable risk. Ssekamatte et al.^17^ demonstrated in a comparable urban Ugandan sample that polysubstance use amplified sexual risk-taking beyond the effects of any individual substance, a finding with direct methodological and interventional implications for the present study population.

### Tramadol as an Undercharacterised High-Risk Substance

The tramadol-specific risk estimates documented here are, to our knowledge, among the highest reported for any substance-RSB association in the West African literature. Tramadol users were 49.6 times more likely to have sex with casual partners, 31.4 times more likely to engage in transactional sex, and 22.5 times more likely to have multiple sexual partners compared to non-users. These odds ratios substantially exceed those for alcohol across the same RSB categories, and the absolute RSB prevalence among tramadol users (88.9% condomless sex, 94.4% casual sex, 77.8% transactional sex) describes a near-universal engagement in high-risk practices within this subgroup.

Alhassan^4,5^ has qualitatively documented tramadol’s social context in Ghana (its use as a perceived masculinity enhancer and a strategy for prolonging sexual performance) but the downstream sexual health consequences had not previously been quantified at this scale. The pharmacological mechanism is plausible: tramadol’s action on mu-opioid receptors and its inhibition of serotonin and norepinephrine reuptake produce a constellation of effects including euphoria, disinhibition, and altered perception of risk and consequence.^10^ These effects are compounded by tramadol’s near-universal co-use with shisha (83.3%) and marijuana (74.1%), creating a polypharmacological environment in which the disinhibitory potential is substantially greater than any single substance alone. Ritchwood et al.^31^ in a meta-analysis of adolescent substance use and RSB, identified polydrug use as a multiplicative rather than additive risk factor (a finding that contextualizes the extraordinarily high odds ratios observed for tramadol users in this study).

### The RSB-STI Gradient and the Architecture of Transmission Risk

The RSB-STI associations (with gonorrhea ORs reaching 36.76 for multiple partners and 29.41 for transactional sex) are broadly consistent with, though in several instances exceed, estimates from comparable sub-Saharan African studies. Seidu et al.^32^ reported an OR of 32.2 between multiple sexual partners and self-reported STIs among Ghanaian men, and Manu et al.^33^ documented high condomless sex-STI associations in a nationally representative Ghanaian sample. The South African data from Kularatne et al.^27^ on gonorrhea and chlamydia prevalence of 10 to 20% in untreated high-risk groups provides a comparison benchmark that validates the magnitude of the present self-reported estimates.

Transactional sex, with HIV positivity of 44.6% among those engaged compared to 8.5% among those who did not, is the single RSB most tightly coupled to adverse STI outcomes in this dataset. Its combination with the high RSB-substance use associations documented in Table 3 [alcohol (OR = 13.30), tramadol (OR = 31.39)] confirms that transactional sex is the primary conduit through which the drug-RSB-STI pathway reaches its most severe clinical endpoint. Kiene and Subramanian^34^ similarly identified alcohol use during sex as the RSB most strongly event-correlated with unprotected intercourse across population-based surveys in sub-Saharan Africa, consistent with the present finding that alcohol use during sex was associated with 40.2% HIV positivity in this sample.

The near-total failure of healthcare engagement (78.3% not seeking treatment and 86.9% not disclosing to partners) transforms what might otherwise be a manageable individual risk into a population-level transmission driver. Kalichman et al.^35^ demonstrated in a Cape Town sample that non-disclosure among HIV-positive individuals receiving STI treatment was a major driver of onward transmission, with partner notification rates below 15% (almost identical to the 13.1% disclosure rate documented in the present study). This convergence across settings suggests that low disclosure is not a culturally idiosyncratic finding but a structural feature of high-stigma, low-service environments that applies directly to Ablekuma North.

## IMPLICATIONS FOR POLICY AND PUBLIC HEALTH PRACTICE

The findings carry direct implications across multiple levels of the Ghanaian public health system. At the national level, tramadol’s disproportionate association with transactional sex (OR = 31.39) and multiple partnerships (OR = 22.49) requires its explicit prioritization within Ghana’s drug control architecture. The National Narcotics Control Commission (NACOC) has historically focused on cannabis and alcohol; the present data provide quantitative evidence base for repositioning opioid misuse (specifically tramadol) as a sexual health concern requiring integration into both drug control and STI prevention programming.

For the Ghana Health Service and Ministry of Health, the central operational implication is the dismantling of the parallel architecture that currently separates substance abuse services from sexual and reproductive health services. The regression evidence demonstrates that drug abuse predicts RSB far more powerfully than any demographic factor; it follows that STI prevention programs that do not address substance use are addressing downstream behavior while ignoring its primary upstream driver. Co-location of STI testing, contraceptive services, and substance use counselling within youth-friendly health facilities (a model tested successfully in combination prevention programs in South Africa and Uganda^17,24^) represents the most evidence-aligned structural response.

The unemployment-transactional sex dynamics demands a multi-sectoral policy response. The co-occurrence of high tertiary education (61.6%) and high unemployment (42%), and their joint association with transactional sex and substance use, implicates economic exclusion as an upstream structural determinant of sexual health risk.^23,29^ Linking youth employment programs to health behavior interventions (an approach recommended by the WHO’s social determinants of health framework) is warranted by the present data. Peer-led prevention models are particularly appropriate given the central role of subjective norms in shaping RSB documented in this and related Ghanaian studies.^18,28^

Finally, the low testing rate (25.6%) and high unawareness of HIV status (59.4%) indicate that the HIV surveillance and testing infrastructure in Ablekuma North is wholly inadequate relative to the epidemic burden documented. Decentralized, community-based testing initiatives (including self-testing) have demonstrated effectiveness in reaching untested populations in comparable West African urban settings and should be prioritized for scale-up in this municipality.

## STRENGTHS AND LIMITATIONS

This study has several notable strengths. The integration of substance use, RSB, and STI outcomes within a single community-based sample provides an internally consistent epidemiological profile of the drug-RSB-STI triad that prior Ghanaian studies have not achieved. The disaggregated, substance-specific analysis (spanning three substances across eight RSBs and five STI outcomes) generates a granularity of evidence directly useful for targeted intervention design. Stratified random sampling across all 14 electoral areas of the municipality ensures proportional geographic representation. The validated instruments demonstrated excellent reliability (α = .927 and .929), and all regression assumptions were met and verified. The motivational data on substance use, which reveal intentional drug use for sexual performance enhancement, adds a dimension of aetiological insight absent from most RSB-focused studies in the region.

Several limitations must be acknowledged. The cross-sectional design precludes causal inference; the temporal sequence from substance use to RSB to STI transmission is inferred from pharmacological and epidemiological reasoning rather than longitudinal demonstration. All STI outcomes rely on self-reported symptoms, which are subject to recall bias, social desirability bias, and the misclassification of asymptomatic infections, particularly important given that 59.4% of participants were unaware of their HIV status, suggesting that a substantial proportion of infections were not captured by symptom reporting. Biological specimen testing would substantially improve outcome validity in future work. The study was confined to Ablekuma North Municipality; generalization to other Ghanaian settings requires caution, although the findings are consistent with patterns from comparable sub-Saharan African urban settings.

## CONCLUSION

Risky sexual behavior is highly prevalent among urban youth in Ablekuma North, spanning condomless sex (50.9%), unplanned sex (47.5%), and transactional sex (19.6%), rates that approach those of traditionally defined high-risk groups. Substance use, particularly tramadol and in the context of pervasive polysubstance co-use, is the primary modifiable determinant of this RSB burden, explaining 40.9% of RSB variance beyond all demographic controls. The drug-RSB-STI pathway operates through specific behavioral conduits (transactional sex and alcohol use during intercourse) that generate the most severe STI risk profiles. That 78.3% of symptomatic participants did not seek treatment, and 86.9% did not disclose to partners, transforms individual infection risk into sustained community-level transmission. Effective response requires integrated, multi-level interventions that simultaneously address substance use, sexual risk behavior, and the structural economic conditions that amplify both in this urban Ghanaian setting.

## Data Availability

All relevant data supporting the findings of this study are included within the manuscript. The dataset collected and/or analyzed during the current study are not publicly available due to the sensitive nature of the data (sexual behavior and substance use) but are available from the corresponding author on reasonable request, subject to ethical approval.

## AUTHOR CONTRIBUTION

- **Conceptualization:** Osei-Mensah Rockson, Faustina Hayford Blankson
- **Methodology:** Osei-Mensah Rockson, Faustina Hayford Blankson
- **Formal analysis:** Osei-Mensah Rockson
- **Investigation:** Osei-Mensah Rockson
- **Data curation:** Osei-Mensah Rockson
- **Writing – original draft:** Osei-Mensah Rockson
- **Writing – review & editing:** Faustina Hayford Blankson
- **Supervision:** Faustina Hayford Blankson

## DATA AVAILABILITY STATEMENT

The dataset collected and/or analyzed during the current study are not publicly available due to the sensitive nature of the data (sexual behavior and substance use) but are available from the corresponding author on reasonable request, subject to ethical approval.

